# A National Genomic Framework for Breast Cancer Risk Stratification in UAE

**DOI:** 10.64898/2026.02.16.26346446

**Authors:** Daniel Matias-Sanchez, Fawad Khan, Rifaat Rawashdeh, Ahmed Alshehhi, Wadha Mohammed Abdurlahman, Aashish Jha, Abdelrahman Saad, Ahmed Al Awadhi, Albarah El-Khani, Andreas Henschel, Asma Al Mannaei, Aziz Khan, Azza Attia, Budour Alkaf, Eduardo da Veiga Beltrame, Fahed Al Marzooqi, Gurunath Katagi, Haiguo Wu, Hakeem Al Mabrazi, Hanil Sajad, Imam Chishty, Joseph Mafofo, Mohamed Alameri, Mohamed El-Hadidi, Omar Soliman, Pierre Zalloua, Raony Cardenas, Senchao Zhang, Shalini Behl, Shilp Purohit, Thyago Cardoso, Val Zvereff, Vinay Kusuma, Wael Elamin, Youseff Idaghdour, Sahar Al Marzooqi, Tiago R Magalhaes, Stephen Grobmyer, Javier Quilez

## Abstract

**Background:** The genetic architecture of Breast Cancer (BC) in Arab populations remains largely understudied, limiting the precision of current prevention and screening programs. The Emirati Genome Program (EGP), one of the world’s first nation-wide sequencing initiatives, offers an unprecedented opportunity to study inherited BC risk across an entire population.

**Methods:** We analyzed 436,780 EGP individuals, including 229,309 women, integrating whole-genome sequencing (WGS) with electronic health records (EHRs). We quantified the prevalence and penetrance of pathogenic and likely pathogenic (P/LP) variants across 13 NCCN-recommended BC genes, evaluated the performance of established polygenic risk scores (PRS), and reconstructed >48,000 pedigrees to measure familial aggregation.

**Results:** P/LP variants were identified in 0.84% of women, accounting for 5.2% of BC cases (mean age of 45.9±11.1 years). Highly penetrant BRCA1 c.4065_4068del (p.Asn1355fs) and BRCA2 c.2808_2811del (p.Ala938Profs) variants showed age-specific cumulative risks of 37.6% and 31% by age 60, respectively, and allele frequencies up to tenfold higher in the Emirati population than in global reference datasets. The European-derived PRS model (PGS000004) demonstrated strong performance, advancing 10-year BC risk onset by a decade for women in the top decile. Family-based PRS discriminated affected from unaffected individuals, revealing higher polygenic risk even within sister pairs.

**Conclusions:** Nation-scale genome sequencing reveals, for the first time, the comprehensive landscape of inherited BC susceptibility within a Middle Eastern population. The integration of monogenic, polygenic, and familial data establishes a national framework for genomic risk stratification, transforming population genomics into a foundation for precision prevention and early detection in the UAE and beyond.

## Introduction

Breast cancer (BC) is the most frequently diagnosed cancer among women worldwide, accounting for ∼24.5% of all female cancer cases and remaining a leading cause of cancer mortality in women^1^. Despite widespread implementation of mammographic, clinical, and genetic screening programs, early detection challenges persist across populations^2^. This is particularly relevant in the Arab Gulf region, where more than one quarter of BC cases are diagnosed before age 40, and the median age at diagnosis is roughly a decade younger than in Western populations^3,4^

Genetic susceptibility to BC arises from both rare high-impact variants in a limited number of genes—most notably *BRCA1* and *BRCA2*—and the combined effect of numerous common variants captured by polygenic risk scores (PRS)^5–7^. Gene-panel testing for high-risk genes has been widely adopted in clinical practice, but it identifies a relatively low number (∼5%) of all BC cases^8^. PRS can complement BC risk prediction and represents a powerful, increasingly validated tool for stratifying BC risk. However, clinical translation remains limited, in part because PRS models – derived primarily from European cohorts – may not generalize well to individuals of other ancestries^9^, an assumption that remains insufficiently tested in large Middle Eastern populations.

The Emirati Genome Program (EGP), a national initiative of the United Arab Emirates (UAE)^10^ aims to perform whole-genome sequencing (WGS) across the entire Emirati population, which comprises approximately 1.3 million citizens. This represents the largest national-scale effort in a non-Western population and, importantly, one of the few national programs approaching complete population coverage^11–13^, enabling nationwide genomics-based public health initiatives. Phenotypic data (Pheno-EGP) are extracted from the electronic health records (EHRs) of the EGP participants through the Malaffi National Health Information Exchange. The nearly complete coverage of the national population, including large highly consanguineous families, makes the EGP a unique resource for human genetics studies and public health initiatives. The EGP’s potential encompasses the full spectrum of human genetic research, from rare disease investigation and the discovery of novel pathogenic variants, as exemplified by recent work on inherited vision loss that reclassified pathogenic variants and uncovered novel findings^14^, to population-scale analyses with direct clinical application, as demonstrated in this manuscript. Together, these complementary approaches position the EGP as a transformative platform for precision medicine, with the power to reshape clinical genomics and deliver tangible health benefits at both individual and population scales.

This work is a nationwide characterization of BC genetic risk integrating conventional high-risk monogenic variant screening with multi-gene risk models as well family and medical information. We (i) quantify the population-wide burden and penetrance of pathogenic variants in commonly screened BC genes; (ii) evaluate the predictive power of BC PRS models both at the population level and within families; and (iii) develop an integrative framework combining rare, common, and familial risk components. These results illustrate the potential of large-scale genomic data to inform precision prevention strategies for BC, though further validation and implementation studies are required to assess clinical impact.

## Methods

### Study Population

We analyzed data from 436,780 Emiratis including 229,309 women, enrolled in the EGP, which integrates WGS data with 10 years longitudinal EHR data from the Abu Dhabi Malaffi Health Information Exchange. BC status was ascertained using SNOMED, ICD-9, and ICD-10 codes. All participants provided informed consent, and this study was approved by the Abu Dhabi Department of Health Medical Research and Development.

### Genomic Sequencing and Quality Control

Participants’ DNA was sequenced at approximately 30× coverage using Illumina NovaSeq 6000 and MGI DNBSEQ platforms. Illumina data were processed with DRAGEN (v3.9–4.1.7)^15^ and MGI data with either Sentieon^16^ or ZBOLT^17^ analysis workflows, in all cases aligned to the GRCh38 reference genome. Quality-control criteria included sequencing yield >87 Gb, mean coverage ≥25.5×, heterozygous-to-homozygous ratio <2.5, estimated contamination <0.05, and mapping rate >75%. After applying these filters, no evidence of platform-specific bias was observed, with variant-call quality and genotype concordance remaining highly consistent across sequencing technologies^18^.

### Variant and Polygenic Risk Analysis

We evaluated 12,453 pathogenic and likely pathogenic (P/LP) variants curated from ClinVar^19^ (≥1-star review status) in the 13 BC susceptibility genes (*ATM, BARD1, BRCA1, BRCA2, CDH1, CHEK2, NF1, PALB2, PTEN, RAD51C, RAD51D, STK11*, and *TP53*) recommended by the National Comprehensive Cancer Network (NCCN)^20^. Carrier status was defined by the presence of any such variant, and its allele frequency (AF) was compared with genome sequencing data from gnomAD 4.0^21^. Age-specific penetrance was estimated using a Kaplan–Meier time-to-event model, where affected carriers contributed age at diagnosis as the event time and unaffected carriers contributed age at last follow-up as censoring time. Cumulative penetrance at each age was defined *as 1−S(t)*, with standard errors and 95% confidence intervals obtained using Greenwood’s formula. Estimates were derived for prespecified ages (30–80 years) for each gene and variant. Ancestry was inferred through principal component analysis (PCA) using PLINK 1.9^22^ and admixture deconvolution using ADMIXTURE^23^ and the populations from the 1000 Genomes Project Phase 3 GRCh38 call set (2022 update, 3,202 samples)^24^. PRS were computed with PRSice-2^25^ using published models developed in European^26^ (PGS000004), East-Asian^27^ (PGS002294), and African^28^ (PGS005104) ancestries and available from the PGS Catalog^29^.

### Family Pedigrees Reconstruction

Pairwise kinship coefficients were estimated from genome-wide SNP data using KING v2.3.2^30^ to reconstruct family pedigrees, including sibling-sibling pairs. These pedigrees were used to assess segregation of pathogenic variants, familial aggregation of PRS, and within-family risk gradients.

### Statistical Analysis

Statistical analyses were performed in Python 3.12 using pandas 2.2.2, NumPy 1.26.4, SciPy 1.15, scikit-learn 1.6.1, and seaborn 0.13.2. Group comparisons used Mann–Whitney U tests, Welch’s t-tests, and binomial tests. Logistic regression models were used to estimate associations between PRS and BC status, reporting odds ratios per SD increase and area under the receiver-operating-characteristic curve (AUC) for discrimination, with 95% confidence intervals derived by bootstrapping. Within-family PRS enrichment was assessed under a null probability of 1/n that the affected woman has the highest PRS among the n female relatives. Significance was derived using a weighted Z-statistic and permutation-based p-values obtained from 200,000 Monte Carlo permutations. Absolute risk curves by PRS percentile were generated using age-stratified logistic regression. Unless otherwise stated, all tests were two-sided, and statistical significance was defined as P < 0.05.

## Results

### Younger Age at Breast Cancer Diagnosis Among Emirati Women

In the 229,309 Emirati women in our study, 1,299 (0.57%) had BC as per the ICD-9, ICD-10, or SNOMED codes in the EHR. Most cases occurred between 45 and 69 years of age (57.7%), with 37.9% clinically diagnosed before 45 years and 4.4% at 70 years or older, compared with 50%, 11.1% and 38.9%, respectively, in European women^31^ (Table S1; Fig. S1). Among the remaining women, 199,404 (87%) were classified as high-confidence controls, and 28,606 were flagged as having benign or ambiguous BC-related records. Table S2 in the Supplementary Appendix details the phenotype codes used for these classifications.

### Population-Wide Penetrance of Pathogenic Variants in Breast Cancer Susceptibility Genes

In the full cohort, 1,929 women (0.84%) carried at least one P/LP variant in the 13 BC-associated genes, representing 363 distinct variants (2.93% of 12,389 total high-risk variants screened). Over 75% of such carriers (n=1,501) harbored variants in *BRCA1, BRCA2, CHEK2*, or *PALB2*, with variants distributed across the genes, without evidence of recurrent hotspots (Fig. S2 in the Supplementary Appendix). Among BC cases carrying a P/LP variant (n=67), 74% involved *BRCA1* or *BRCA2*. Consistent with this predominance, Kaplan–Meier estimates showed marked age-dependent penetrance for these genes, with *BRCA1* carriers reaching a cumulative risk of 22.6% by age 50 and rising to 46.9% by age 70; and *BRCA2* carriers showing 13.9% risk by age 50 and 35.6% by age 70. *PALB2* and *CHEK2* contributed an additional 20% of affected carriers, with substantially lower cumulative risks overall: penetrance reached 4.3% for *PALB2* and 3.5% for *CHEK2* by age 50, rising to 13.0% and 8.7%, respectively, by age 70. Pathogenic variants in *ATM, TP53*, and *RAD51D* were rare among cases (<1% each), with modest age-specific risks (generally 1–10% depending on age). No BC cases were observed among carriers of *RAD51C, BARD1, CDH1*, or *STK11* (Table S3).

Among the 45 variants with nonzero penetrance estimates (Table 1), highly penetrant and prevalent alleles included NM_007294.4(*BRCA1*):c.4065_4068del(p.Asn1355fs) (n=92) and *NM_000059*.*3(BRCA2):c*.*2808_2811del (p*.*Ala938Profs)* (n=37). Age-stratified analyses showed a steep increase in penetrance for both alleles, with *BRCA1* penetrance rising from 12.4% at by age 40 to 29.8% at by age 50, 37.6% by age 60, and 68.8% by age 70. The risk allele in *BRCA2* followed a similar pattern, increasing from 5.3% by age 40 to 17.2% by age 50, 31.0% by age 60, and 48.3% by age 70 (Figure 1). These BRCA1 and BRCA2 alleles together accounted for nearly 30% of all monogenic BC cases in the cohort, with allele frequencies approximately six- and ten-fold higher in Emiratis than in global reference populations in gnomAD^21^. In contrast, the variant-specific alleles NM_024675.4(*PALB2*):c.3350+4A>G (n=251) and NM_007194.4(*CHEK2*):c.409C>T (p.Arg137Ter) (n=117)—although more frequent—showed near-zero penetrance at younger ages, with risk emerging only later in life. The *PALB2* c.3350+4A>G allele reached a cumulative penetrance of 6.7% by age 60, whereas the *CHEK2* p.Arg137Ter allele showed no measurable penetrance until age 70, at which point penetrance reached 16.7% (Figure1, Table 1).

**Table 1.**
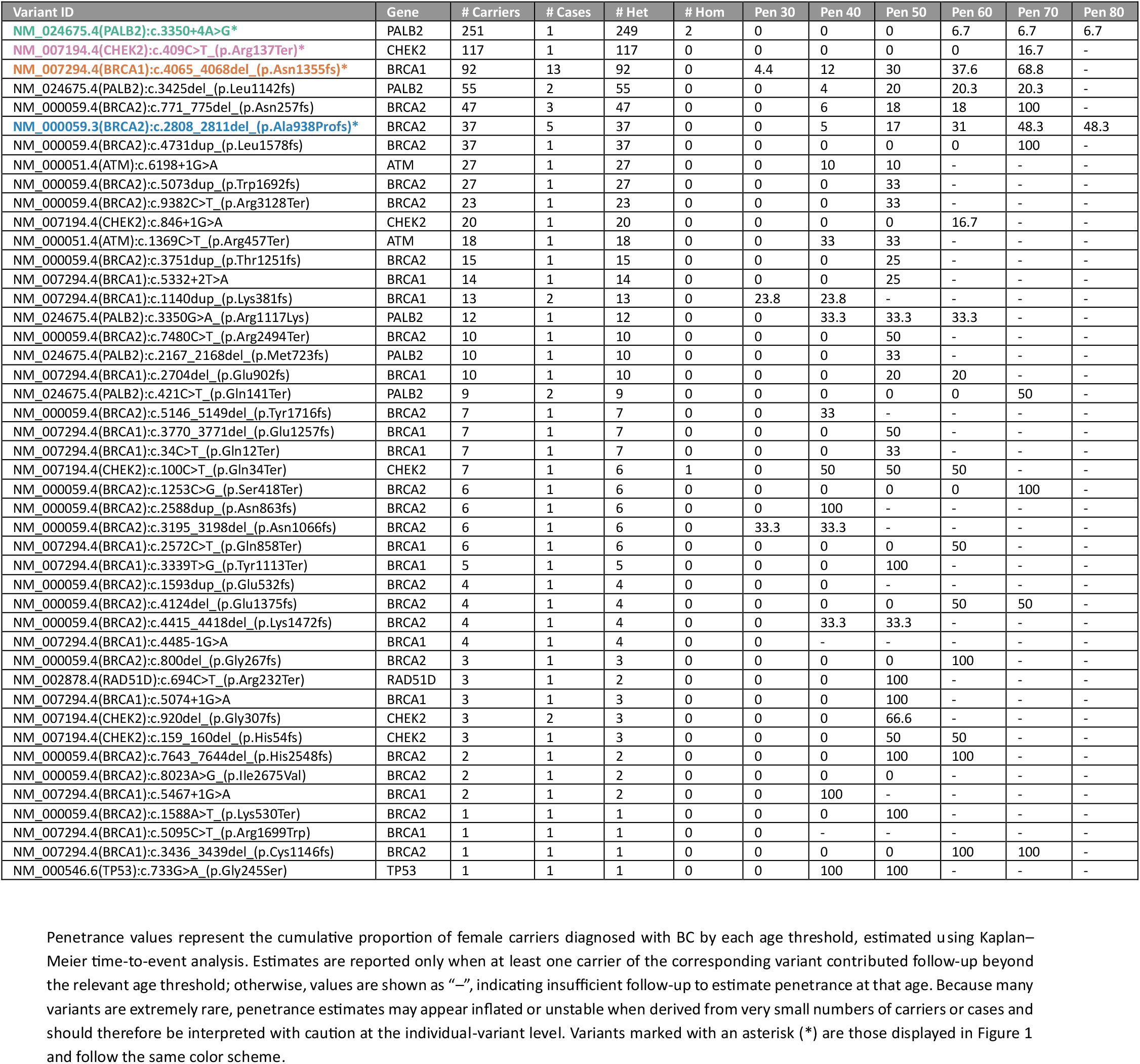
Age-Specific Cumulative Penetrance of Pathogenic and Likely Pathogenic Variants in Breast Cancer Susceptibility Genes.

| Variant ID | Gene | # Carriers | # Cases | # Het | # Hom | Pen 30 | Pen 40 | Pen 50 | Pen 60 | Pen 70 | Pen 80 |
| --- | --- | --- | --- | --- | --- | --- | --- | --- | --- | --- | --- |
| NM_024675.4(PALB2):c.3350+4A>G* | PALB2 | 251 | 1 | 249 | 2 | 0 | 0 | 0 | 6.7 | 6.7 | 6.7 |
| NM_007194.4(CHEK2):c.409C>T (p.Arg137Ter)* | CHEK2 | 117 | 1 | 117 | 0 | 0 | 0 | 0 | 0 | 16.7 | - |
| NM_007294.4(BRCA1):c.4065_4068del (p.Asn1355fs)* | BRCA1 | 92 | 13 | 92 | 0 | 4.4 | 12 | 30 | 37.6 | 68.8 | - |
| NM_024675.4(PALB2):c.3425del (p.Leu1142fs) | PALB2 | 55 | 2 | 55 | 0 | 0 | 4 | 20 | 20.3 | 20.3 | - |
| NM_000059.4(BRCA2):c.771_775del (p.Asn257fs) | BRCA2 | 47 | 3 | 47 | 0 | 0 | 6 | 18 | 18 | 100 | - |
| NM_000059.3(BRCA2):c.2808_2811del (p.Ala938Profs)* | BRCA2 | 37 | 5 | 37 | 0 | 0 | 5 | 17 | 31 | 48.3 | 48.3 |
| NM_000059.4(BRCA2):c.4731dup (p.Leu1578fs) | BRCA2 | 37 | 1 | 37 | 0 | 0 | 0 | 0 | 0 | 100 | - |
| NM_000051.4(ATM):c.6198+1G>A | ATM | 27 | 1 | 27 | 0 | 0 | 10 | 10 | - | - | - |
| NM_000059.4(BRCA2):c.5073dup (p.Trp1692fs) | BRCA2 | 27 | 1 | 27 | 0 | 0 | 0 | 33 | - | - | - |
| NM_000059.4(BRCA2):c.9382C>T (p.Arg3128Ter) | BRCA2 | 23 | 1 | 23 | 0 | 0 | 0 | 33 | - | - | - |
| NM_007194.4(CHEK2):c.846+1G>A | CHEK2 | 20 | 1 | 20 | 0 | 0 | 0 | 0 | 16.7 | - | - |
| NM_000051.4(ATM):c.1369C>T (p.Arg457Ter) | ATM | 18 | 1 | 18 | 0 | 0 | 33 | 33 | - | - | - |
| NM_000059.4(BRCA2):c.3751dup (p.Thr1251fs) | BRCA2 | 15 | 1 | 15 | 0 | 0 | 0 | 25 | - | - | - |
| NM_007294.4(BRCA1):c.5332+2T>A | BRCA1 | 14 | 1 | 14 | 0 | 0 | 0 | 25 | - | - | - |
| NM_007294.4(BRCA1):c.1140dup (p.Lys381fs) | BRCA1 | 13 | 2 | 13 | 0 | 23.8 | 23.8 | - | - | - | - |
| NM_024675.4(PALB2):c.3350G>A (p.Arg1117Lys) | PALB2 | 12 | 1 | 12 | 0 | 0 | 33.3 | 33.3 | 33.3 | - | - |
| NM_000059.4(BRCA2):c.7480C>T (p.Arg2494Ter) | BRCA2 | 10 | 1 | 10 | 0 | 0 | 0 | 50 | - | - | - |
| NM_024675.4(PALB2):c.2167_2168del (p.Met723fs) | PALB2 | 10 | 1 | 10 | 0 | 0 | 0 | 33 | - | - | - |
| NM_007294.4(BRCA1):c.2704del (p.Glu902fs) | BRCA1 | 10 | 1 | 10 | 0 | 0 | 0 | 20 | 20 | - | - |
| NM_024675.4(PALB2):c.421C>T (p.Gln141Ter) | PALB2 | 9 | 2 | 9 | 0 | 0 | 0 | 0 | 0 | 50 | - |
| NM_000059.4(BRCA2):c.5146_5149del (p.Tyr1716fs) | BRCA2 | 7 | 1 | 7 | 0 | 0 | 33 | - | - | - | - |
| NM_007294.4(BRCA1):c.3770_3771del (p.Glu1257fs) | BRCA1 | 7 | 1 | 7 | 0 | 0 | 0 | 50 | - | - | - |
| NM_007294.4(BRCA1):c.34C>T (p.Gln12Ter) | BRCA1 | 7 | 1 | 7 | 0 | 0 | 0 | 33 | - | - | - |
| NM_007194.4(CHEK2):c.100C>T (p.Gln34Ter) | CHEK2 | 7 | 1 | 6 | 1 | 0 | 50 | 50 | 50 | - | - |
| NM_000059.4(BRCA2):c.1253C>G (p.Ser418Ter) | BRCA2 | 6 | 1 | 6 | 0 | 0 | 0 | 0 | 0 | 100 | - |
| NM_000059.4(BRCA2):c.2588dup (p.Asn863fs) | BRCA2 | 6 | 1 | 6 | 0 | 0 | 100 | - | - | - | - |
| NM_000059.4(BRCA2):c.3195_3198del (p.Asn1066fs) | BRCA2 | 6 | 1 | 6 | 0 | 33.3 | 33.3 | - | - | - | - |
| NM_007294.4(BRCA1):c.2572C>T (p.Gln858Ter) | BRCA1 | 6 | 1 | 6 | 0 | 0 | 0 | 0 | 50 | - | - |
| NM_007294.4(BRCA1):c.3339T>G (p.Tyr1113Ter) | BRCA1 | 5 | 1 | 5 | 0 | 0 | 0 | 100 | - | - | - |
| NM_000059.4(BRCA2):c.1593dup (p.Glu532fs) | BRCA2 | 4 | 1 | 4 | 0 | 0 | 0 | - | - | - | - |
| NM_000059.4(BRCA2):c.4124del (p.Glu1375fs) | BRCA2 | 4 | 1 | 4 | 0 | 0 | 0 | 0 | 50 | 50 | - |
| NM_000059.4(BRCA2):c.4415_4418del (p.Lys1472fs) | BRCA2 | 4 | 1 | 4 | 0 | 0 | 33.3 | 33.3 | - | - | - |
| NM_007294.4(BRCA1):c.4485-1G>A | BRCA1 | 4 | 1 | 4 | 0 | 0 | - | - | - | - | - |
| NM_000059.4(BRCA2):c.800del (p.Gly267fs) | BRCA2 | 3 | 1 | 3 | 0 | 0 | 0 | 0 | 100 | - | - |
| NM_002878.4(RAD51D):c.694C>T (p.Arg232Ter) | RAD51D | 3 | 1 | 2 | 0 | 0 | 0 | 100 | - | - | - |
| NM_007294.4(BRCA1):c.5074+1G>A | BRCA1 | 3 | 1 | 3 | 0 | 0 | 0 | 100 | - | - | - |
| NM_007194.4(CHEK2):c.920del (p.Gly307fs) | CHEK2 | 3 | 2 | 3 | 0 | 0 | 0 | 66.6 | - | - | - |
| NM_007194.4(CHEK2):c.159_160del (p.His54fs) | CHEK2 | 3 | 1 | 3 | 0 | 0 | 0 | 50 | 50 | - | - |
| NM_000059.4(BRCA2):c.7643_7644del (p.His2548fs) | BRCA2 | 2 | 1 | 2 | 0 | 0 | 0 | 100 | 100 | - | - |
| NM_000059.4(BRCA2):c.8023A>G (p.Ile2675Val) | BRCA2 | 2 | 1 | 2 | 0 | 0 | 0 | 0 | - | - | - |
| NM_007294.4(BRCA1):c.5467+1G>A | BRCA1 | 2 | 1 | 2 | 0 | 0 | 100 | - | - | - | - |
| NM_000059.4(BRCA2):c.1588A>T (p.Lys530Ter) | BRCA2 | 1 | 1 | 1 | 0 | 0 | 0 | 100 | - | - | - |
| NM_007294.4(BRCA1):c.5095C>T (p.Arg1699Trp) | BRCA1 | 1 | 1 | 1 | 0 | 0 | - | - | - | - | - |
| NM_007294.4(BRCA1):c.3436_3439del (p.Cys1146fs) | BRCA2 | 1 | 1 | 1 | 0 | 0 | 0 | 0 | 100 | 100 | - |
| NM_000546.6(TP53):c.733G>A (p.Gly245Ser) | TP53 | 1 | 1 | 1 | 0 | 0 | 100 | 100 | - | - | - |

**Figure 1.**
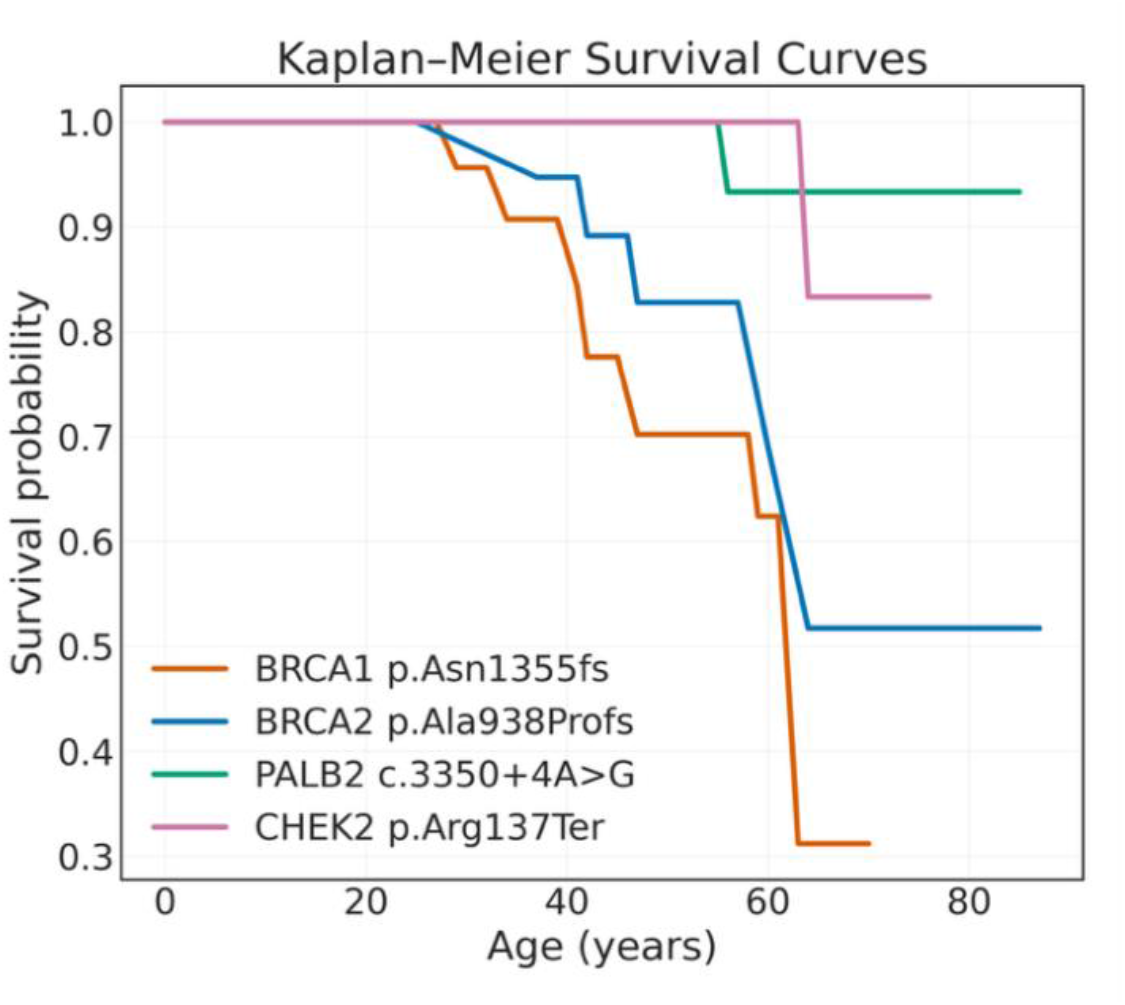
Age-Specific Cumulative Penetrance of Breast Cancer for Pathogenic Variants in *BRCA1, BRCA2, PALB2*, and *CHEK2*. Kaplan–Meier estimates of age-specific cumulative BC penetrance are shown for carriers of *BRCA1* p.Asn1355fs, *BRCA2* p.Ala938Profs, *PALB2* c.3350+4A>G, and *CHEK2* p.Arg137Ter. Individuals without a BC diagnosis were censored at their last known age. Differences in curve shape illustrate gene- and variant-specific variation in age-dependent BC risk.

Nevertheless, only 5.2% of the 1,299 women with clinically diagnosed BC carried a P/LP variant in these genes, indicating that monogenic factors explain just a small fraction of the total burden of disease^8^. This observation highlights the need for complementary approaches, such as PRS, to capture the broader heritable component of susceptibility.

### Polygenic Risk Modeling in the Emirati Population

We evaluated three established PRS models for BC developed in European (PGS000004)^26^, East Asian (PGS002294)^27^, and African (PGS005104)^28^ cohorts. The European-derived model showed the highest predictive performance in our Emirati cohort with an AUC of 0.63 (95% CI, 0.60–0.65), closely matching the performance reported in the original work (AUC = 0.63). The East Asian and African models performed worse in our cohort, with AUCs of 0.60 and 0.57, respectively, compared to their originally reported values of 0.639 and 0.60 (Fig. S3 in the Supplementary Appendix). Consequently, we used the European-derived PRS model for all subsequent analyses. These results are consistent with population-structure analyses from ADMIXTURE, showing that Emirati individuals clustered most closely with European reference populations, with an average genetic similarity of ∼70%, followed by South Asian (≈18%) and African (≈11%) components (Fig. S4 in the Supplementary Appendix).

### Polygenic Score Stratifies the Timing of Breast Cancer Risk

To translate PRS into clinically actionable risk estimates, we calculated 10-year absolute BC risk across PRS percentiles (Figure 2). Women in the top decile (≥90th percentile) reached a 10-year risk of approximately 2.8% by age 50, a level observed in the overall population only around age 60, indicating an advancement of risk by roughly a decade. Conversely, women in the lowest quartile (≤25th percentile) reached a comparable 10-year risk (∼1.6%) only by age 60, about 10 years later than average. At ages 40, 50, and 60, the 10-year BC risk for high-PRS (90th percentile) women reaches approximately 0.8%, 2.9% and 5.7%, corresponding to ∼2.6-, 2.9- and 3.6-fold higher risk compared with low-PRS (25th percentile) women.

**Figure 2.**
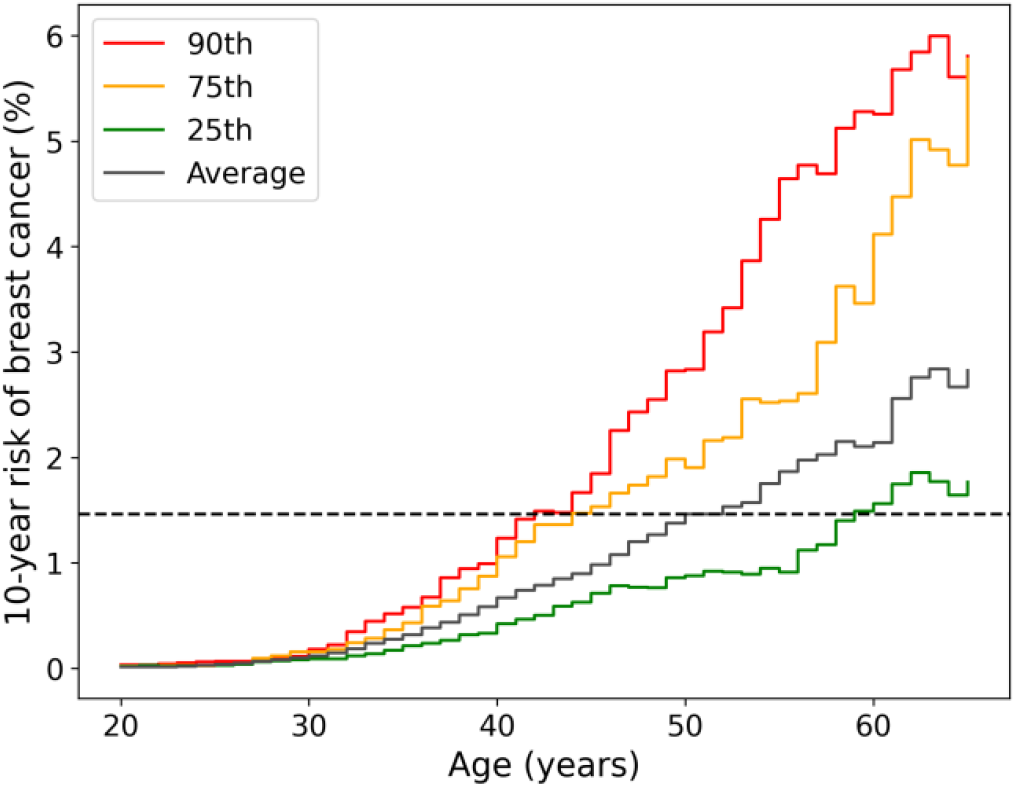
Age-Specific 10-Year Risk of BC by Polygenic Risk Percentile. Women were stratified by PRS percentile groups (≥90th, ≥75th, ≤25th percentiles, and population average), and age-specific 10-year absolute BC risk was estimated. Risk curves show a clear PRS-dependent gradient, with higher PRS associated with earlier and steeper increases in absolute risk across age. Women in the highest PRS decile reached population-average risk levels approximately 10 years earlier than average-risk women, whereas those in the lowest PRS quartile showed delayed risk accumulation. The dashed line indicates the cohort-average 10-year BC risk at age 50, illustrating both risk acceleration and delay across PRS strata.

### Combined Effects of Monogenic and Polygenic Risk

We stratified women by carrier status for *BRCA1, BRCA2, PALB2*, and *CHEK2* and by PRS level, categorized as low (deciles 1–3), intermediate (4–7), or high (8–10). Due to insufficient case numbers, *PALB2* and *CHEK2* did not meet criteria for reliable estimation and were excluded from the final analysis. Among non-monogenic carriers, PRS was strongly associated with BC status (Figure 3). In *BRCA1* carriers, risk was uniformly high across PRS strata, indicating that the penetrance of *BRCA1* mutations is largely independent of polygenic background. By contrast, *BRCA2* carriers exhibited a modest gradient, with higher risk estimates in the upper PRS category.

**Figure 3.**
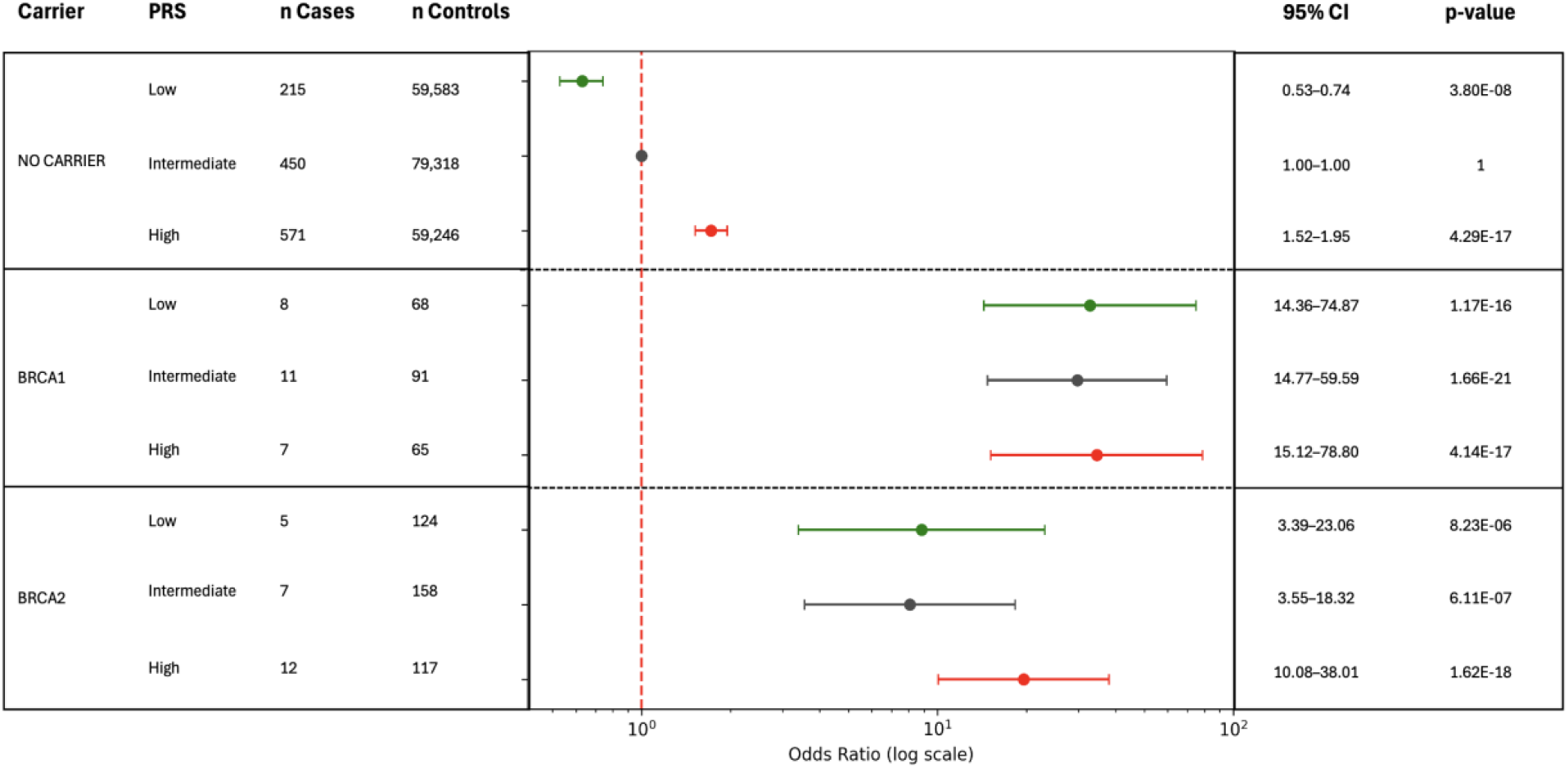
Odds ratios for BC by polygenic risk score (PRS) strata in carriers and non-carriers of pathogenic variants in *BRCA1, BRCA2*. Women were grouped into PRS strata (low, intermediate, high). Relative to non-carrier intermediates, non-carriers showed clear PRS–risk gradients (lower in low, higher in high). *BRCA1* carriers had consistently high risks regardless of PRS, while in *BRCA2* higher PRS tended to increase risk, though estimates were imprecise due to limited power.

### Familial and Polygenic Dimensions of Breast Cancer Risk

We reconstructed 48,289 multi-generational family pedigrees with at least 2 individuals^14^—the largest pedigree resource ever derived from a single population. Among these, 1,086 families included at least one carrier of a P/LP variant in the 13 BC genes screened, providing a concrete framework for family-level clinical surveillance.

Across the cohort, 930 families contained at least one BC case, of which 886 (95.3%) were simplex (only one affected individual) and 44 (4.7%) were multiplex with ≥2 affected relatives. Median age at diagnosis showed only a modest difference between simplex and multiplex cases (48.0 vs. 45.5 years; P = 1.4×10^−2^). Thus, the markedly young age at onset observed in Emirati women cannot be attributed to familial clustering but instead reflects population-wide characteristics rather than concentration of early-onset disease in a small number of high-risk pedigrees.

To assess the contribution of polygenic background to familial clustering, we focused on 47,203 families in which no member carried a monogenic pathogenic variant. Unaffected relatives of BC cases exhibited a markedly higher mean PRS than members of cancer-free families (0.58 vs 0.46; P = 4.9×10^−12^), demonstrating robust aggregation of polygenic burden within families (Fig. S5 in the Supplementary Appendix). Within families containing both affected and unaffected female relatives, BC cases were substantially more likely to exhibit the highest PRS among all women in the family (212/696 informative families; P = 1.45×10^−4^, weighted permutation test; Fig. S6 in the Supplementary Appendix), providing direct evidence that PRS enables individual-level risk discrimination even within shared familial genetic backgrounds, a distinction that is typically not observable in smaller studies

This pattern was confirmed in pairs of sisters in which only one had BC. In 1,210 of such pairs, the affected sister had a higher PRS (58.3%; P = 4.9×10^−9^), with a mean within-pair difference of 0.12 SD units (P = 4.8×10^−11^) (Figure 4). These results demonstrate that PRS retains robust discriminatory power at the individual level even among first-degree relatives, providing direct evidence that polygenic risk can refine risk stratification and inform early detection strategies within families already under clinical surveillance.

**Figure 4.**
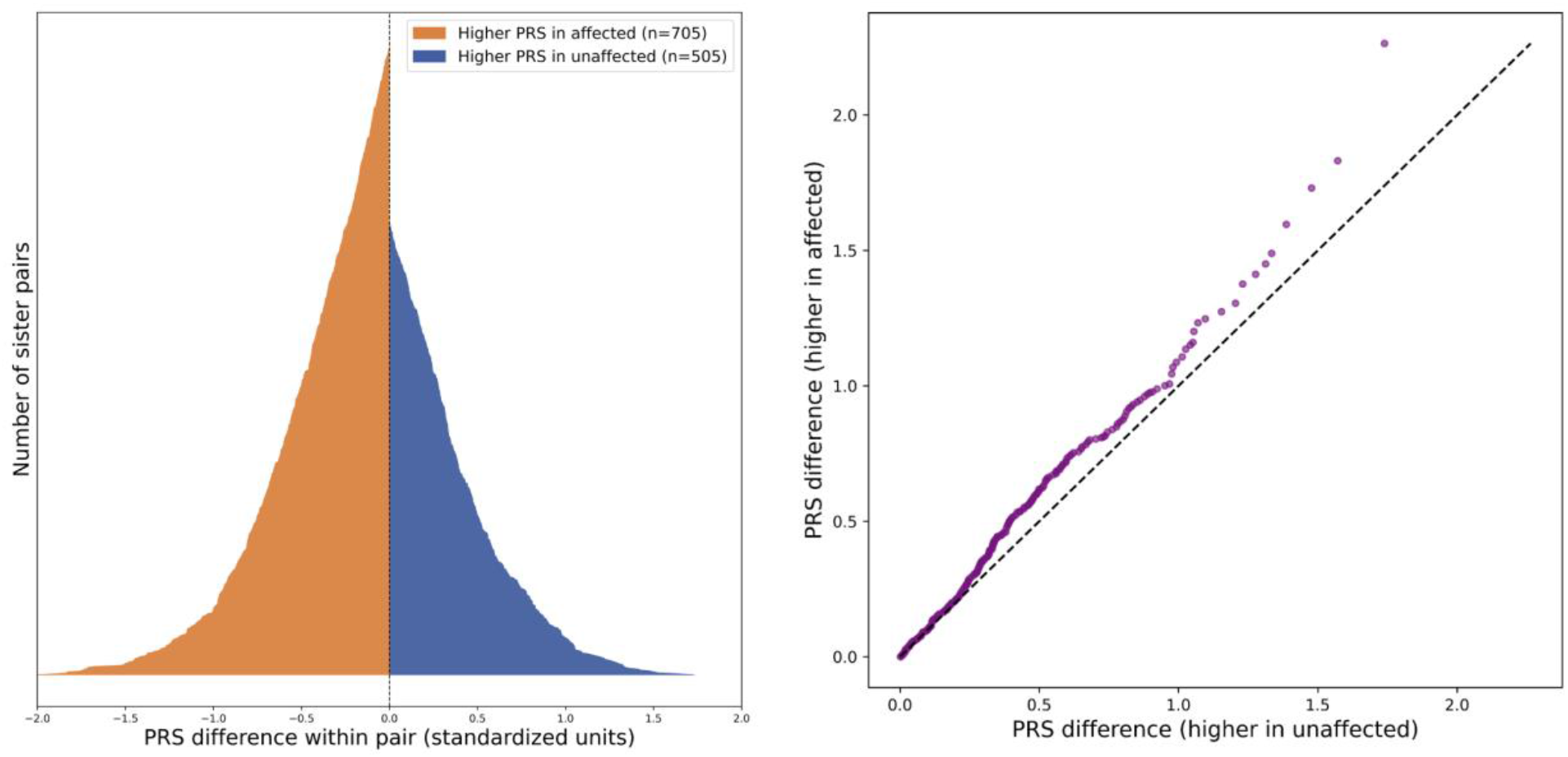
Within-Pair Polygenic-Risk Differences in Breast Cancer–Discordant Sister Pairs. Left Panel shows the distribution of standardized within-pair PRS differences across 1,210 sister pairs discordant for BC, sorted by ΔPRS. Each pair consists of one affected and one unaffected sister; curves are color-coded according to which sibling carried the higher score (orange = affected, blue = unaffected). Affected sisters carried the higher PRS in 705 of 1,210 pairs (58.3%; binomial P = 4.92 × 10^−9^), with a mean within-pair difference of 0.12 standardized units (SD = 0.608; P = 4.79 × 10^−11^). Right panel shows a quantile–quantile comparison of within-pair PRS differences between pairs where the affected sibling had the higher PRS (y-axis) and those where the unaffected sibling had the higher PRS (x-axis). Deviations above the diagonal indicate a right-shifted distribution among affected-higher pairs, consistent with greater PRS separation; formal tests confirmed these differences (Mann–Whitney U P = 2.62 × 10^−3^; Kolmogorov–Smirnov P = 5.72 × 10^−4^).

### Integrated Framework for Genomic Risk Stratification

Together, these analyses support clinical guidelines for BC risk management at a national scale (Table 2). The set of 45 pathogenic variants with demonstrable penetrance captures the monogenic BC cases across the population, with high-penetrant genes *BRCA1 and BRCA2* conferring a 12-fold risk and moderate-penetrant genes *PALB2 and CHEK2* a 3-fold risk, highlighting this set of 45 variants as the clinically relevant pathogenic backbone of hereditary BC.

**Table 2.** Combined Genetic and Familial Contributors to Breast Cancer Risk.

| Risk Category | Odds Ratio (95% CI) | P Value | # Carriers | # Cases | # Controls | Interpretation |
| --- | --- | --- | --- | --- | --- | --- |
| High-penetrant genes (BRCA1, BRCA2) | 12.65 (9.4-16.9) | $2.05 \times 10^{-36}$ | 769 | 50 | 719 | Strongly pathogenic; ~12.7-fold increased risk |
| Moderate-penetrant genes (PALB2, CHEK2) | 3.23 (1.8-5.6) | $3.15 \times 10^{-4}$ | 723 | 13 | 710 | Moderate effect; ~3.1-fold increased risk |
| Low-penetrant genes (Other 9 BC genes) | 1.5 (0.6-4.1) | 0.33 | 459 | 4 | 455 | Low impact; ~1.5-fold increased risk |
| Top 10% PRS (non-monogenic) | 2.26 (1.87-2.49) | $1.52 \times 10^{-22}$ | 22,732 | 237 | 22,495 | High polygenic burden; ~2.3-fold increased risk |
| Family history (non-monogenic) | 1.83 (1.51-2.21) | $7.39 \times 10^{-9}$ | 12,629 | 119 | 12,510 | Familial aggregation; ~1.8-fold increased risk |
| PRS top 10% + Family history | 3.99 (2.63-6.06) | $6.49 \times 10^{-8}$ | 1,095 | 23 | 1,072 | Joint polygenic-familial effect; ~4.0-fold increased risk |

Odds ratios and two-sided P values were obtained from 2x2 contingency analyses (Fisher's exact test), without covariate adjustment. Gene-level categories were defined as BRCA1 and BRCA2 (high penetrance), PALB2 and CHEK2 (moderate penetrance), and all remaining screened genes (ATM, BARD1, CDH1, NF1, PTEN, RAD51C, RAD51D, STK11, TP53) as low penetrance. Counts represent the total number of women carrying at least one pathogenic or likely pathogenic variant in each gene group; "non-monogenic" excludes all such carriers. Polygenic-risk strata (e.g., top 10% PRS) were based on PRS percentiles computed across the full sequenced cohort and restricted to non-monogenic individuals. Family history indicated that a woman had at least one close family member with breast cancer, based on genetically inferred family relationships. This definition captures the presence of an affected first-degree relative. The combined category ("PRS top 10% + family history") includes women who meet both criteria.

Beyond rare variants, polygenic risk adds a powerful and independent layer: women in the top 10% of the PRS distribution show a ∼2.3-fold higher risk. Women with BC family history had ∼1.8 times higher risk to develop BC compared to those without such history – BC family history defined as at least one genetically inferred first-degree relative diagnosed with BC. When high PRS and family history co-occur, risk approaches a four-fold elevation, higher than that of moderate-penetrance gene carriers. Importantly, women in the highest PRS stratum also develop BC roughly a decade earlier than those with average or low scores, indicating that polygenic background not only quantifies lifetime susceptibility but also shifts the timing of risk onset.

Altogether, the EGP provides an integrated framework to identify women currently free of disease who carry measurable genomic predisposition and may benefit from earlier detection or preventive intervention, representing the first demonstration of population-wide genomic risk stratification at national scale.

## Discussion

The EGP provides one of the most comprehensive genomic mappings of BC inherited risk to date within a national population. By integrating (i) rare variant analysis, (ii) PRS, and (iii) family-level information at national scale, we take a crucial step toward resolving the full spectrum of hereditary susceptibility within a single country—transforming population genomics into a foundation for precision public health.

Most clinical work on the use of germline genetic testing to date has focused on the management of individuals. The current study provides a powerful framework for managing both individuals and populations at risk for breast cancer through the implementation of a comprehensive national germline genomic strategy. It highlights the importance of clinical-grade sequencing at the population level, as well as the value of integration with a national electronic medical record system.

At present, data from the EGP are being used to manage individual patients who have or are at risk for breast cancer, as well as their families. Programs such as the one described in this study are currently under development to enable the pre-emptive identification of large cohorts of individuals prior to the onset of cancer, and to support risk stratification aimed at implementing a more personalized and cost-effective breast cancer screening strategy for the country^32^. Insights and opportunities for population-level risk modification and disease management are uniquely enabled by the EGP, not only for breast cancer but also for other heritable cancers in the future.

The clinical implications are substantial. Conventional genetic testing identifies only a small fraction of affected women through a handful of high-penetrance variants^33,34^. With nationwide sequencing, these alleles can now be comprehensively catalogued, and their carriers identified, enabling empirical, age-specific estimation of penetrance. Equally transformative is the potential to use PRS clinically at population scale to capture the broader heritable gradient of risk, identifying women who would remain invisible to current clinical screening. Unlike traditional population studies^35–37^, EGP aims to sequence the entire national population—meaning that percentile thresholds reflect true population-wide distributions, not estimates based on sampling or imputation.

We further demonstrate that polygenic scores trained in European populations are transferable and clinically meaningful in Arab ancestry, retaining predictive value both across the population and within families. PRS not only distinguishes high- and low-risk women but also discriminates between close relatives, even identifying the sibling most likely to develop disease. This shows that polygenic information can complement monogenic screening^38,39^ and family history to enable individualized, genetically informed prevention at both population and familial levels.

At the national scale, integrating these three layers—monogenic, polygenic, and familial—creates a unified framework for precision prevention. In this framework, all carriers can be identified through systematic national sequencing; every family with an affected member can be traced; and every high-polygenic-risk individual can be prioritized for enhanced surveillance^40^. Implemented through public-health systems, such a framework could transform BC control from reactive detection to proactive, data-driven prevention, offering earlier screening, tailored interventions, and cascade testing at unprecedented scale. This work, conducted by a multidisciplinary collaboration of scientists, clinicians, and public health regulators, underscores the clinical applicability of this approach and its potential to inform precision prevention strategies directly within national healthcare systems.

Challenges remain. Many participants are below the age of peak incidence, and long-term follow-up will refine these risk estimates. Current pathogenic annotations still rely heavily on ClinVar and genome-wide association studies, which remain inherently biased toward variants identified in patients of European descent^41,42^. This underscores the need to reclassify variants of uncertain significance and to identify Arab-specific pathogenic alleles through continued national sequencing efforts^43–45^. Integrating environmental and lifestyle data with genomic predictors will also be essential to fully model disease risk^46,47^.

In summary, the EGP illustrates how sequencing an entire nation enables a complete, quantitative understanding of inherited cancer risk—free from sampling bias and externally defined thresholds. It provides a practical blueprint for next-generation screening programs worldwide: a model where every citizen’s genome contributes to prevention, and where precision oncology begins not in the clinic, but across the nation.

## Data Availability

All data produced in the present study are available upon reasonable request to the Department of Health, Abu Dhabi

