## Supplementary Appendix for "A National Genomic Framework for Breast Cancer Risk Stratification in UAE"

Public

**Supplementary Table 1. Age-stratified incidence and prevalence of breast cancer in Emirati women compared to European Commission data.**

| Age | # Samples EGP | # Cases EGP | Prevalence EGP | Incidence EGP | European commission incidence |
|---|---|---|---|---|---|
| 0-19 | 65,722 | 10 | 0.02 | 0.8 | 0 |
| 20-44 | 108,891 | 482 | 0.44 | 37.1 | 11.1 |
| 45-69 | 48,085 | 750 | 1.56 | 57.7 | 50 |
| >=70 | 6,611 | 57 | 0.86 | 4.4 | 38.9 |

Breast cancer cases in the EGP cohort (n = 229,309) were stratified by age group and compared to incidence rates reported by the European Commission. Percentages reflect the proportion of total cases observed within each age category.

**Supplementary Table 2. Breast cancer phenotype definitions used to classify cases and moderate-confidence controls based on ICD9/10 and SNOMED codes.**

| Cohort type | Disease Codes | Phenotype | Number of Individuals in 230K Cohort |
|---|---|---|---|
| Cases | ICD10: C50 (& subtypes) | Malignant neoplasm of breast | 1261 |
| | ICD10: D05 (& subtypes) | Carcinoma in situ of breast | 249 |
| | ICD9: 174 | Malignant neoplasm of female breast | 366 |
| | ICD9: 175 | Malignant neoplasm of male breast | 2 |
| | ICD9: 233 | Carcinoma in situ of breast and genitourinary system | 42 |
| | SNOMED: 254837009 | Malignant tumor of breast | 65 |
| Moderate-Confidence Controls | ICD10: Z15.01 | Genetic susceptibility to malignant neoplasm of breast | 70 |
| | ICD10: Z86.000 | Personal history of in-situ neoplasm of breast | 14 |
| | ICD10: Z85.3 | Personal history of malignant neoplasm of breast | 257 |
| | ICD10: Z80.3 | Family history of malignant neoplasm of breast | 3612 |
| | ICD10: Z42.1 | Encounter for breast reconstruction following mastectomy | 20 |
| | ICD10: Z12.31 | Encounter for screening mammogram for malignant neoplasm of breast | 23798 |
| | ICD10: Z12.39 | Encounter for other screening for malignant neoplasm of breast | 13947 |
| | ICD10: Z12.3 | Encounter for screening for malignant neoplasm of breast | 26140 |
| | ICD10: R92.0 | Mammographic microcalcification found on diagnostic imaging of breast | 543 |
| | ICD10: R92.1 | Mammographic calcification found on diagnostic imaging of breast | 243 |
| | ICD10: R92.2 | Inconclusive mammogram | 2215 |
| | ICD10: R92.8 | Other abnormal and inconclusive findings on diagnostic imaging of breast | 7723 |
| | ICD10: R92 | Abnormal and inconclusive findings on diagnostic imaging of breast | 8935 |
| | ICD10: D03.52 | Melanoma in situ of breast (skin) (soft tissue) | 0 |
| | ICD10: D49.3 | Neoplasm of unspecified behavior of breast | 71 |
| | ICD10: D48.6 | Neoplasm of uncertain behavior of breast | 406 |
| | ICD10: D24.1 | Benign neoplasm of right breast | 1964 |
| | ICD10: D24.2 | Benign neoplasm of left breast | 1729 |
| | ICD10: D24.9 | Benign neoplasm of unspecified breast | 1195 |
| | ICD10: D24 | Benign neoplasm of breast | 3126 |
| | ICD10: C79.81 | Secondary malignant neoplasm of breast | 3 |
| | ICD10: C44.591 | Other specified malignant neoplasm of skin of breast | 0 |
| | ICD10: C44.501 | Unspecified malignant neoplasm of skin of breast | 4 |
| | ICD10: C44.511 | Basal cell carcinoma of skin of breast | 1 |
| | ICD10: C4A.52 | Merkel cell carcinoma of skin of breast | 0 |

* One individual can present more than one ICD10 code. If a sample has a Case disease code, is not considered for moderate-cofidence control disease codes

Public

Public

### Supplementary Tabe 3. Age-specific cumulative penetrance for breast cancer among female carriers of pathogenic/likely pathogenic variants in monogenic breast cancer genes (Kaplan–Meier estimates).

| Gene | nVariants in Clinvar | nVariants in EGP F | nCarriers F | Het | Hom | nCases | Penetrance 30 yo | Penetrance 40 yo | Penetrance 50 yo | Penetrance 60 yo | Penetrance 70 yo | Penetrance 80 yo |
|---|---|---|---|---|---|---|---|---|---|---|---|---|
| ATM | 1,671 | 63 | 222 | 221 | 1 | 2 | 0 | 2.5±1.5 | 2.5±1.5 | 2.5±1.5 | 2.5±1.5 | - |
| BARD1 | 451 | 22 | 87 | 87 | 0 | 0 | 0 | 0 | 0 | 0 | 0 | 0 |
| BRCA1 | 3,321 | 67 | 302 | 302 | 0 | 26 | 2.6±1.3 | 7.7±2.4 | 22.6±4.6 | 33.6±7.2 | 46.9±10.2 | - |
| BRCA2 | 4,343 | 113 | 474 | 474 | 0 | 24 | 0.4±0.4 | 3.7±1.5 | 13.9±3.4 | 21.2±4.7 | 35.6±7.8 | 35.6±7.8 |
| CDH1 | 180 | 9 | 13 | 13 | 0 | 0 | 0 | - | 0 | - | - | - |
| CHEK2 | 764 | 29 | 316 | 313 | 3 | 6 | 0 | 0.7±0.5 | 3.5±1.7 | 4.9±2.2 | 8.7±4.1 | 8.7±4.1 |
| NF1 | 1 | 0 | 0 | 0 | 0 | 0 | - | - | - | - | - | - |
| PALB2 | 1,091 | 25 | 409 | 407 | 2 | 7 | 0 | 1.2±0.7 | 4.3±2.2 | 7.6±3.8 | 13±6.5 | 42±18.7 |
| PTEN | 16 | 1 | 1 | 1 | 0 | 0 | 0 | 0 | - | - | - | - |
| RAD51C | 182 | 7 | 76 | 76 | 0 | 0 | 0 | 0 | 0 | 0 | 0 | 0 |
| RAD51D | 175 | 10 | 38 | 38 | 0 | 1 | 0 | 0 | 9.1±6.8 | 9.1±6.8 | 9.1±6.8 | - |
| STK11 | 133 | 4 | 4 | 4 | 0 | 0 | 0 | 0 | 0 | 0 | 0 | 0 |
| TP53 | 125 | 14 | 24 | 24 | 0 | 1 | 0 | 9.1±6.8 | 9.1±6.8 | 9.1±6.8 | 9.1±6.8 | 9.1±6.8 |

Cumulative penetrance values represent the estimated probability (in %) of developing breast cancer before a given age among female carriers of pathogenic/likely pathogenic variants. Estimates were derived using Kaplan–Meier survival analysis, where time-to-event was age at diagnosis for cases and age at last follow-up for controls. Standard errors (SE) were calculated using Greenwood's formula. "–" indicates insufficient follow-up to estimate penetrance at that age.

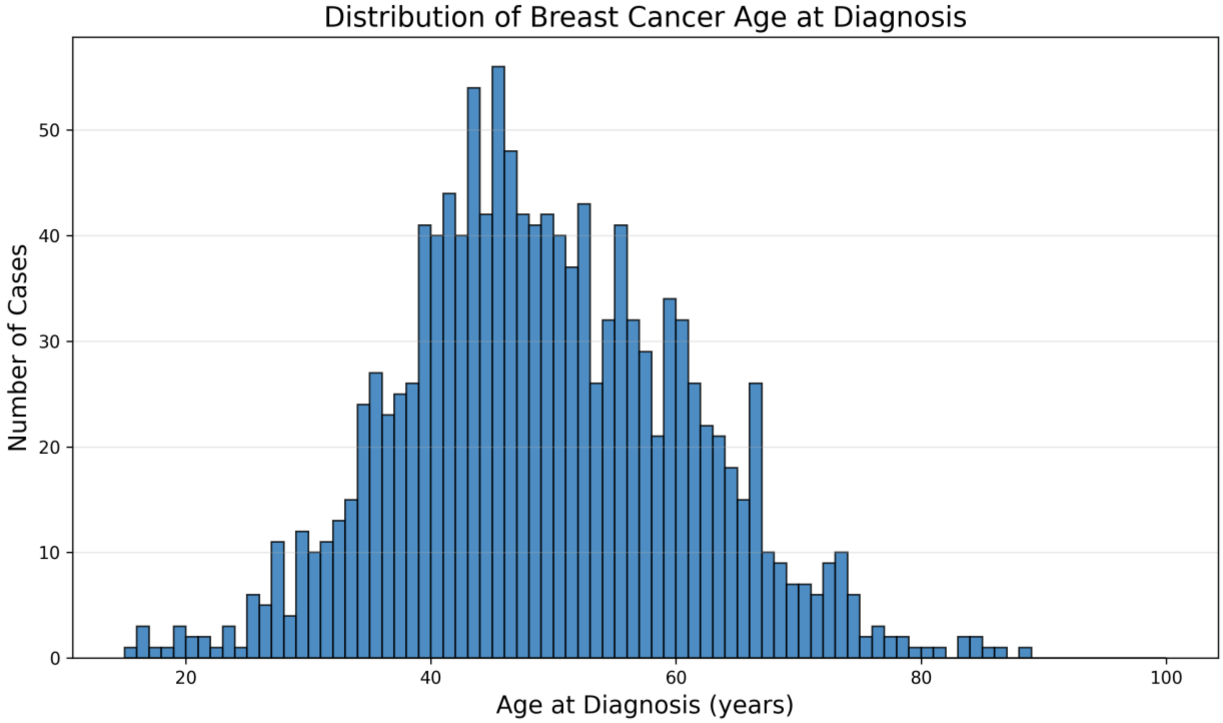


**Supplementary Figure 1. Distribution of Age at Breast Cancer Diagnosis Among Emirati Women.** Shown is the histogram of age at diagnosis for all women with breast cancer in the Emirati Genome Program. Each bar represents the number of cases diagnosed at a given age (in one-year increments).

Public

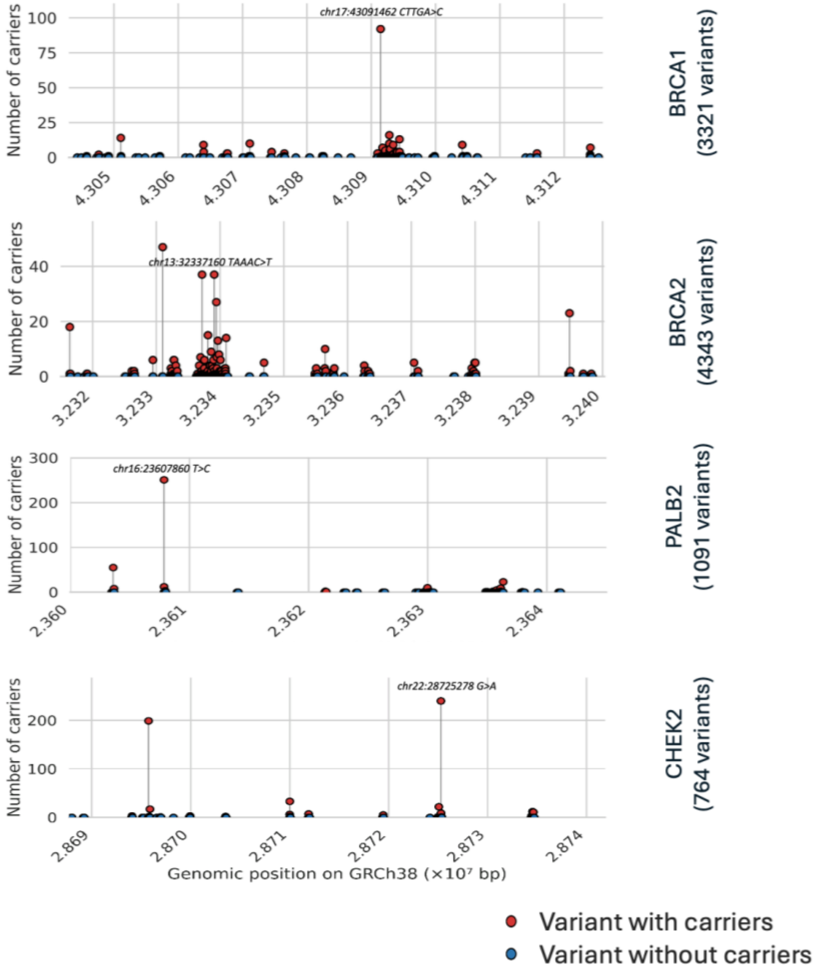


**Supplementary Figure 2. Lolliplots of pathogenic variants in three breast-cancer genes** (BRCA1: 3321 variants and 302 carriers; BRCA2: 4343 variants and 474 carriers; PALB2: 1091 variants and 409 carriers and CHEK2: 764 variants and 316 carriers) showing the number of carriers per variant in the EGP (red: variants with ≥1 carrier; blue: zero carriers)

Public

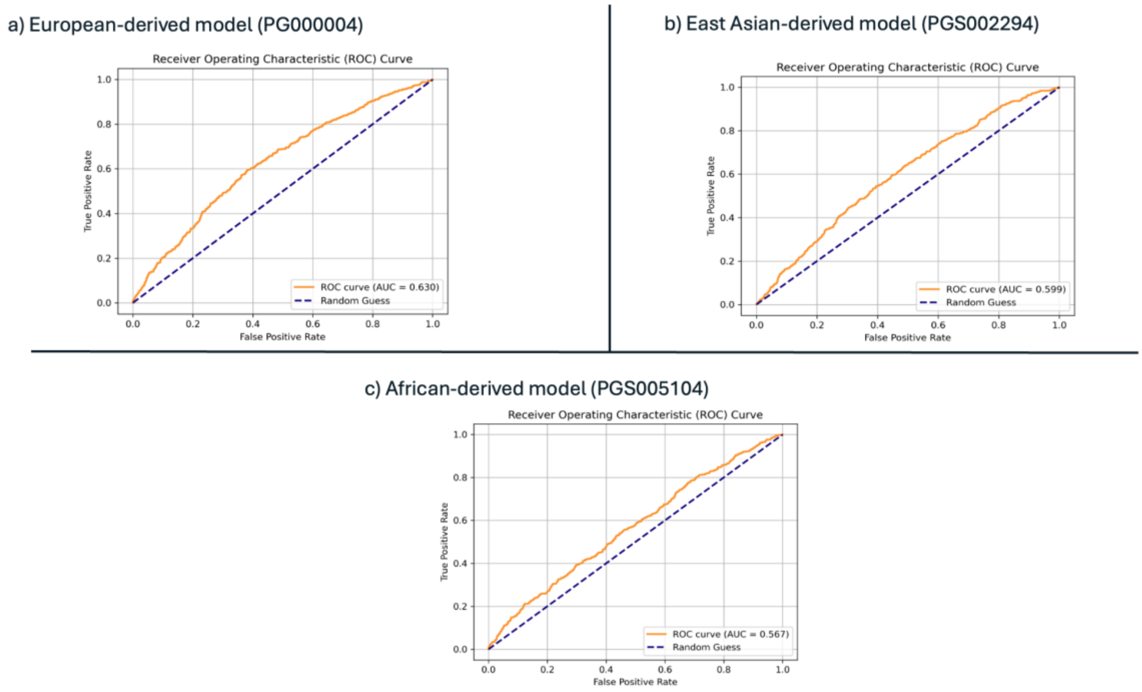


**Supplementary Figure 3. Performance of breast cancer polygenic risk score (PRS) models in the Emirati Genome Program cohort.** Receiver Operating Characteristic (ROC) curves are shown for three published PRS models: (a) European-derived model (PGS000004, AUC = 0.630), (b) East Asian-derived model (PGS002294, AUC = 0.599), and (c) African-derived model (PGS005104, AUC = 0.567). The European-derived model demonstrated the highest discrimination in Emirati women, followed by the East Asian- and African-derived models, although all models showed reduced predictive accuracy compared to performance in their discovery populations.

Public

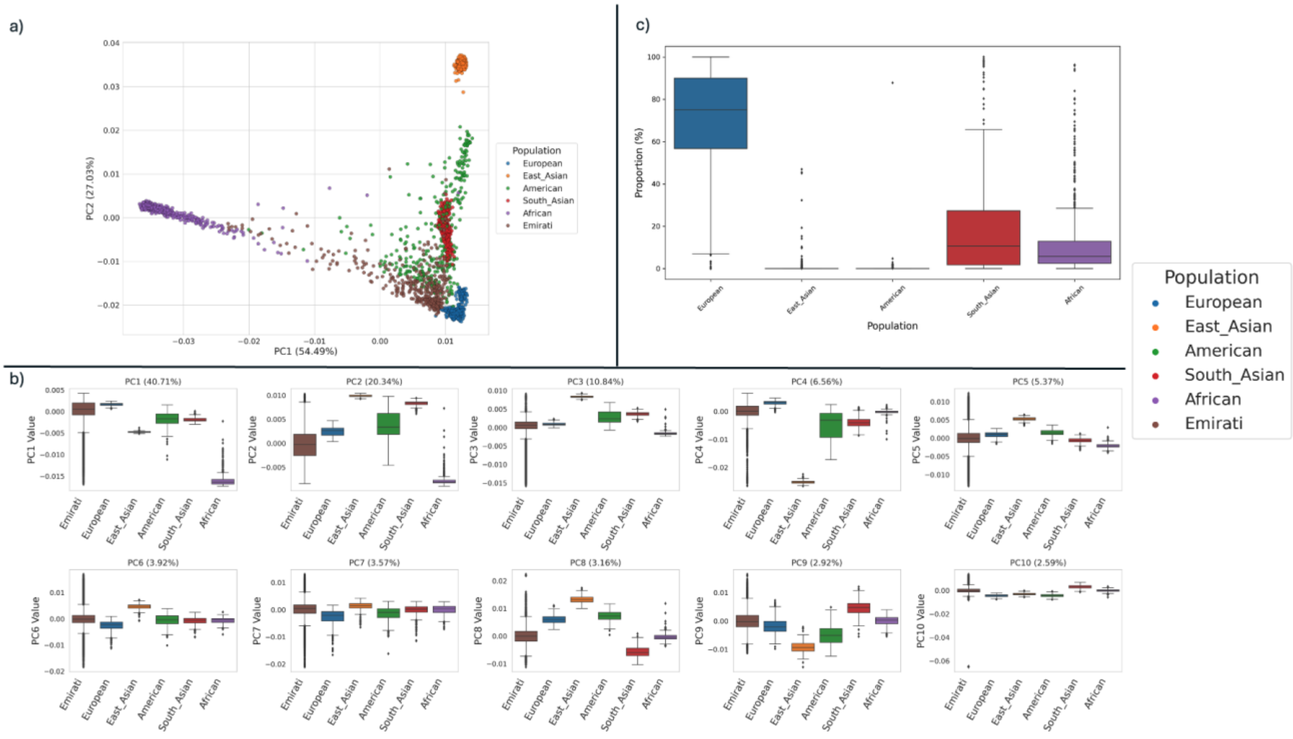


**Supplementary Figure 4. Ancestry analysis of the Emirati cohort by comparison to 1000 Genomes superpopulations. (a)** Principal component analysis of a subset of 400 Emirati women from the 230K cohort combined with 2 503 reference individuals from the 1000 Genomes Project (1KGP). PC1 (x-axis; 54.49% variance) separates African versus non-African populations, while PC2 (y-axis; 27.03% variance) defines a European–South Asian continuum. Emirati samples (brown) span a broad admixed range, clustering between European (blue), South Asian (red) and African (purple) references and distant from East Asian (orange) groups. **(b)** Distributions of PCs 1–10 by superpopulation. Boxplots summarize the spread of each principal component within each 1KGP superpopulation, recapitulating the major axes of genetic variation seen in (a). **(c)** Supervised admixture proportions for 1 000 randomly sampled Emirati individuals relative to the five 1KGP superpopulations. Each boxplot shows the distribution of ancestry fractions (y-axis) for European, East Asian, American, South Asian, and African components (x-axis), demonstrating predominant European contribution with secondary South Asian and African admixture.

Public

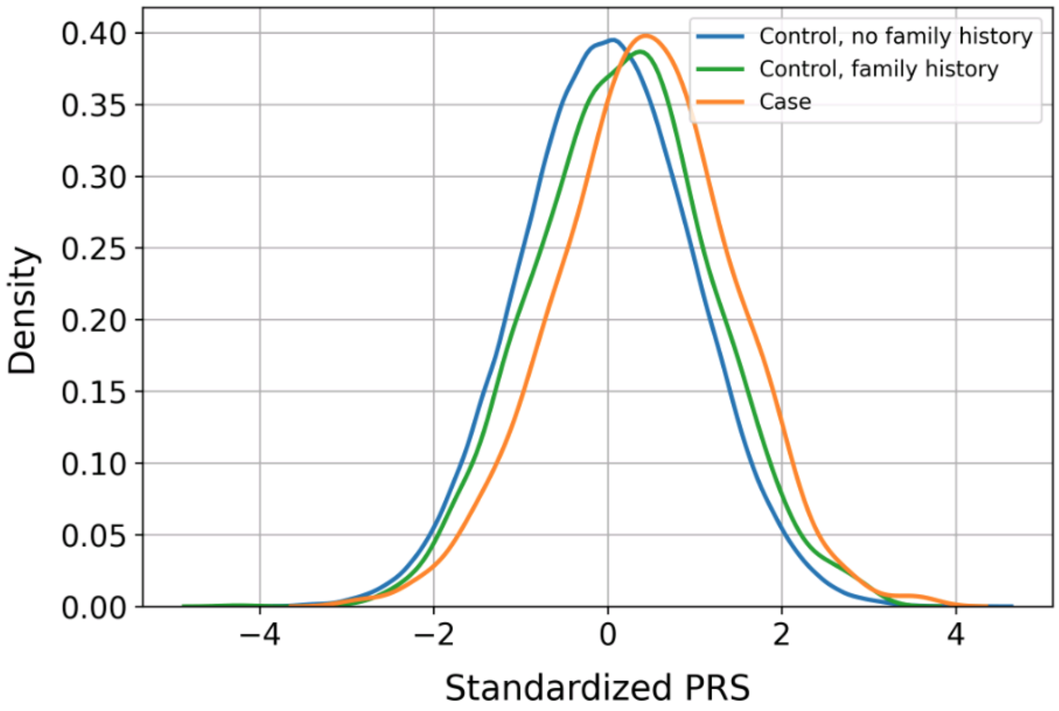


**Supplementary Figure 5. Family-history evaluation of PRS enrichment.** PRS in monogenic−negative controls with versus without a first-degree affected relative. Among 1,985 monogenic-negative controls with a first-degree family history compared to 196,786 with no such history, mean standardized PRS was significantly elevated (0.224 vs. –0.005; Mann–Whitney $P < 1\times10^{-3}$).

Public

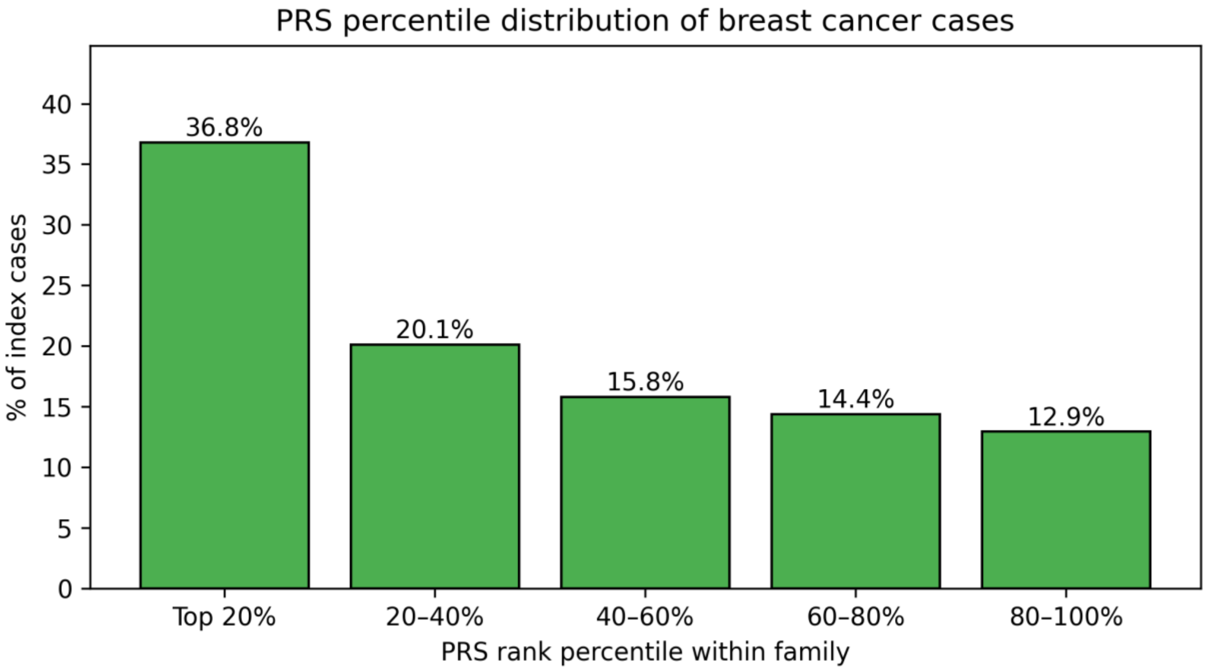


**Supplementary Figure 6. Distribution of Breast-Cancer Index Cases Across Polygenic-Risk Percentiles Within Families.** PRS rank of the index case in families with one affected woman. In clusters with a single breast cancer case, we show the proportion of cases ranking 1st (highest PRS) to 5th or lower within their family. 40.1% ranked 1st, significantly more than expected by chance ($P = 2.98 \times 10^{-4}$)

Public
